# NOVEL PLASMA GLYCOPROTEIN BIOMARKERS PREDICT PROGRESSION-FREE SURVIVAL IN SURGICALLY RESECTED CLEAR CELL RENAL CELL CARCINOMA

**DOI:** 10.1101/2022.01.11.21268517

**Authors:** Daniel Serie, Amanda A. Myers, Daniela A. Haehn, Alexander S. Parker, Essa Bajalia, Giovanni Gonzalez, Qiongyu Li, Maurice Yu Wong, Kaitlynn Moser, Bo Zhou, David D. Thiel

**Affiliations:** InterVenn Biosciences; Mayo Clinic Florida Department of Urology; University of Florida, College of Medicine - Jacksonville

**Keywords:** Renal cell carcinoma, glycoproteins, biomarker, progression free survival

## Abstract

**Introduction:** Limited data exists on utilization of protein post-translational modifications as biomarkers for clear cell renal cell carcinoma (ccRCC). We employed high-throughput glycoproteomics to evaluate differential expression of glycoprotein-isoforms as novel markers for ccRCC progression-free survival (PFS).

**Methods:** Plasma samples were obtained from 77 patients treated surgically for ccRCC. Glycoproteomic analyses were carried out after liquid chromatography tandem mass spectrometry. Age-adjusted Cox proportional hazard models were constructed to evaluate PFS. Optimized Harrells c-index was employed to dichotomize the collective for the construction of Kaplan-Meier curves.

**Results:** The average length of follow-up was 3.4 (range: 0.04-9.83) years. Glycoproteomic analysis identified 39 glycopeptides and 14 non-glycosylated peptides that showed statistically significant (false discovery rate p ≤0.05) differential expression associated with PFS. Five of the glycosylated peptides conferred continuous hazard ratio of > 6 (range 6.3-11.6). These included prothrombin A2G2S glycan motif (HR=6.47, P=9.53E-05), immunoglobulin J chain FA2G2S2 motif (HR=10.69, P=0.001), clusterin A2G2 motif (HR=7.38, P=0.002), complement component C8A A2G2S2 motif (HR=11.59, P=0.002), and apolipoprotein M glycopeptide with non-fucosylated and non-sialylated hybrid-type glycan (HR=6.30, P=0.003). Kaplan-Meier curves based on dichotomous expression of these five glycopeptides resulted in hazard ratios of 3.9-10.7, all with p-value < 0.03. Kaplan-Meyer plot using the multivariable model comprising 3 of the markers yielded HR of 11.96 (p <0.0001).

**Conclusion:** Differential glyco-isoform abundance of plasma proteins may be a useful source of biomarkers for the clinical course and prognosis of ccRCC.

## Introduction

The standard of care for clinically localized clear cell renal cell carcinoma (ccRCC) is partial or radical nephrectomy. Distant metastatic disease develops in 20-30% of patients with initially localized disease, despite surgical resection of the primary tumor[1]. Current surveillance strategies for ccRCC recurrence rely primarily on stage-based risk assessment to determine intervals of monitoring between 3 and 12 months. Methods to precisely risk stratify patients represents a clear unmet need in patients with ccRCC. Non-invasive risk stratification tests have potential to guide clinical decision making on follow up. Moreover, in the era of targeted molecular therapeutics, there is an urgent need for improved prognostication in ccRCC patients who would potentially benefit from adjuvant therapy. While several potential biomarkers have been explored in this context, none have demonstrated superiority to pathologic stage [2].

Protein glycosylation is the most common form of post-translational protein modification (PTM) and has profound effects on protein structure, conformation, and function. Changes in PTM are a well-recognized hallmark of malignant transformation, observed in a variety of cancers, and have been associated with severity and progression of disease [3]. While the majority of clinically utilized protein biomarkers (PSA, CEA, or CA125) are glycosylated [4], currently used immuno-tests for these markers only recognize their peptide backbone but do not consider their glycoprotein-isoforms. Early studies evaluating glycoproteins showed promise in the development of biomarkers for various cancers [5, 6], the technical challenges of accurately measuring individual glyco-isoforms have so far precluded clinical use. Recent development of a highly accurate and scalable platform has now made it possible to carry out rapid and accurate analysis of site-specific changes in glycan motifs of multiple serum glycopeptides[7]. We deployed this platform to evaluate whether differential expression of glycosylated peptides may provide insights into the variable natural history and disease course of ccRCC.

## Methods

### 2.1 Sample Procurement

Plasma samples obtained from patients with newly diagnosed ccRCC prior to their surgical treatment, and stored at -80°C, were provided by the Mayo Clinic Renal Registry, an IRB-approved biorepository and clinical database on ccRCC patients. Histopathologic diagnosis of ccRCC was confirmed for all patients by a genitourinary pathologist who served as an independent reviewer, as previously described [8].

### 2.2 Sample Preparation

Serum samples were reduced with dithiothreitol (DTT), alkylated with iodoacetamide (IAA)then digested with trypsin at 37°C for 18 hours. To quench the digestion, formic acid was added to each sample after incubation to a final concentration of 1% (v/v).

### 2.3 LC-MS Analysis

Digested serum samples were separated over an Agilent ZORBAX Eclipse Plus C18 column (2.1 mm x 150 mm i.d., 1.8 µm particle size) using an Agilent 1290 Infinity UHPLC system. The mobile phase A consisted of 3% acetonitrile, 0.1% formic acid in water (v/v), and the mobile phase B of 90% acetonitrile 0.1% formic acid in water (v/v), with the flow rate set at 0.5 mL/minute. The binary solvent composition was set at 100% mobile phase A at the beginning of the run, linearly shifting to 20% B at 20 minutes, 30% B at 40 minutes, and 44% B at 47 minutes. The column was flushed with 100% B and equilibrated with 100% A for a total run time of 70 minutes. After electrospray ionization (ESI), samples were injected into Agilent 6495B triple quadrupole mass spectrometer operated in dynamic multiple reaction monitoring (dMRM) mode. The MRM transitions comprised of 408 glycopeptides. Then normalized by comparing them with the abundance of 68 non-glycosylated peptide fragments from which the glycopeptides monitored were derived, as previously published [7].

### 2.4 Data Analyses

PeakBoundaryNet, an in-house-developed spectrogram feature recognition and integration software based on recurrent neural networks [9] was used to integrate chromatogram peaks and obtain molecular abundance quantification for each peptide and glycopeptide. To account for differential protein abundance, the ratio of glycosylated peptides to non-glycosylated peptides (“relative abundance”) was utilized. To correct for assay-related artifacts, pooled human serum sample was interspersed every ten samples. The glycosylated/non-glycosylated peptide ratios in these calibration samples were used to normalize the patient sample ratios.

The association of relative abundance ratio values with progression-free survival was assessed using age-adjusted Cox Proportional Hazards models. P-values for multiple comparisons were determined using the Benjamini-Hochberg false discovery rate (FDR) [10] procedure. P-value of 0.05 or less was defined as statistically significant. Biomarker expression values were dichotomized at cutoff values that maximized Harrell’s C-index. Kaplan-Meier plots were generated to assess the differential progression-free survival between groups with high and low marker expression. Multivariable predictive models were generated from glycopeptides with HR > 6 and FDR < 0.05, by trimming features via backwards selection, then generating models applied to each sample as a holdout via leave-one-out cross-validation (LOOCV). All statistical analyses were run in R, version 3.4.4.

### 2.5 Ingenuity Pathway Analysis

Bioinformatic analysis was performed to identify canonical pathways and associated protein networks by using Ingenuity® Pathway Analysis software (QIAGEN Inc.). Right-tailed Fisher’s exact test was used to calculate a *P* value of overlap to determine the significance of each canonical pathway, and P <0.01 was considered significant. Ingenuity® Knowledge Base was used to predict the upstream regulators of candidate biomarkers. The molecular filter was applied to analysis transcription factors, cytokines, transmembrane and G-coupled protein receptors. Associated networks with candidate biomarkers were algorithmically generated.

## Results

We analyzed plasma samples of 77 ccRCC patients, 48 (62%) were male and 29 were female (38%). Mean age was 61 years (range: 33-79 years). Fifty-four patients (70%) had stage I disease, 9 (11.7%) stage II, 12 (15.6%) stage III, 1 (1.3%) had stage IV disease, and 1 was missing stage. The average length of follow-up was 3.4 (range: 0.04 – 9.83) years. No patients with T1 or T2 disease had positive margin. Four patients with pT3 disease had positive renal vein margins, of these two had disease progression. In total, thirteen patients suffered recurrent disease (Table 1). The sites of progression were 7 (54%) lung, 2 (15%) bone and 4 (31%) local.

**Table 1.** Patient Characteristics

| Variable | Summary (N=77) |
| --- | --- |
| Age at diagnosis, years | mean 61, range 33-79 |
| Sex, n (%) |  |
| Female | 29 (38%) |
| Male | 48 (62%) |
| Surgery Performed |  |
| Partial Nephrectomy | 30 (39%) |
| Laparoscopic | 19 |
| Open | 11 |
| Radical Nephrectomy | 47 (61%) |
| Laparoscopic | 34 |
| Open | 13 |
| Pathologic Stage |  |
| T1 | 54 |
| T2 | 9 |
| T3 | 12 |
| T4 | 1 |
| Preoperative eGFR, ml/min/1.73m<sup>2</sup> | 79 (65, 92) |
| Furhman Grade |  |
| 1 | 7 |
| 2 | 43 |
| 3 | 20 |
| 4 | 7 |
| Disease Progression | 13 (16%) |

Glycoproteomic analysis identified 39 glycopeptides and 14 non-glycosylated peptides (Appendix A), that showed statistically significant (false discovery rate p≤0.05) differential expression among patients with shorter and longer PFS, respectively. Five of the glycosylated peptides conferred a continuous hazard ratio (HR) of > 6 (range 6.3-11.6). These included the prothrombin A2G2S glycan motif (HR=6.47, P=<0.001), immunoglobulin J chain FA2G2S2 motif (HR=10.69, P=0.001), clusterin A2G2 motif (HR=7.38, P=0.002), complement component C8A A2G2S2 motif (HR=11.59, P=0.002), and apolipoprotein M glycopeptide with non-fucosylated and non-sialylated hybrid-type glycan (HR=6.30, P=0.003). Kaplan-Meier (KM) curves based on dichotomous expression of these five glycopeptides into high score and low score resulted in HR of 3.9-10.7, all with p-value < 0.03 (Figure 1). The median difference between high score and low score was 1.07 for THRB_A2G2S glycan motif, 0.46 for IGJ_FA2G2S2, 0.82 for CLUS_A2G2 motif, 0.86 for CO8A_A2G2S2, and 0.58 for APOM_Hybrid (Figure 1). Three glycoproteins had statistically significant correlation with increasing stage. Prothrombin A2G2S glycan motif (Linear estimate =0.37, P=<0.001), clusterin A2G2 motif (Linear estimate= 0.15, P=0.004), and complement component C8A A2G2S2 motif (Linear estimate = 0.17, P=0.04) associated with increasing tumor stage (Appendix B). Figure 2 represents other glycoprotein markers with continuous HR under 6, but dichotomization resulted in KM curves with HR > 10.

**Figure 1.**
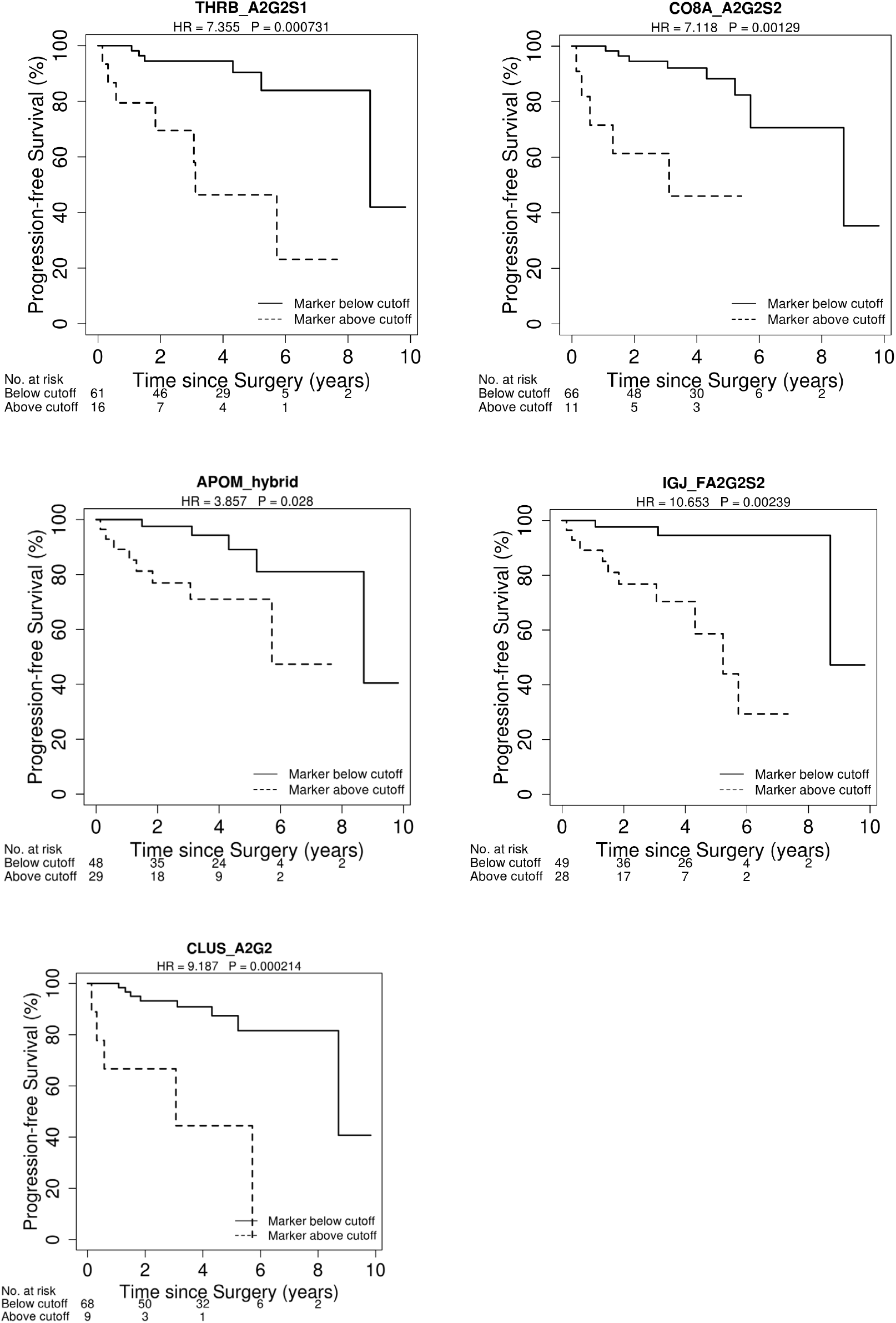
Kaplan Meier curve of Glycoprotein markers utilizing Harrell’s C index to optimize cutoff values. This figure demonstrates the five glycoprotein markers with continuous hazard ratio >6 (resulting in dichotomized hazard ratios from 3.9-10.7).

**Figure 2.**
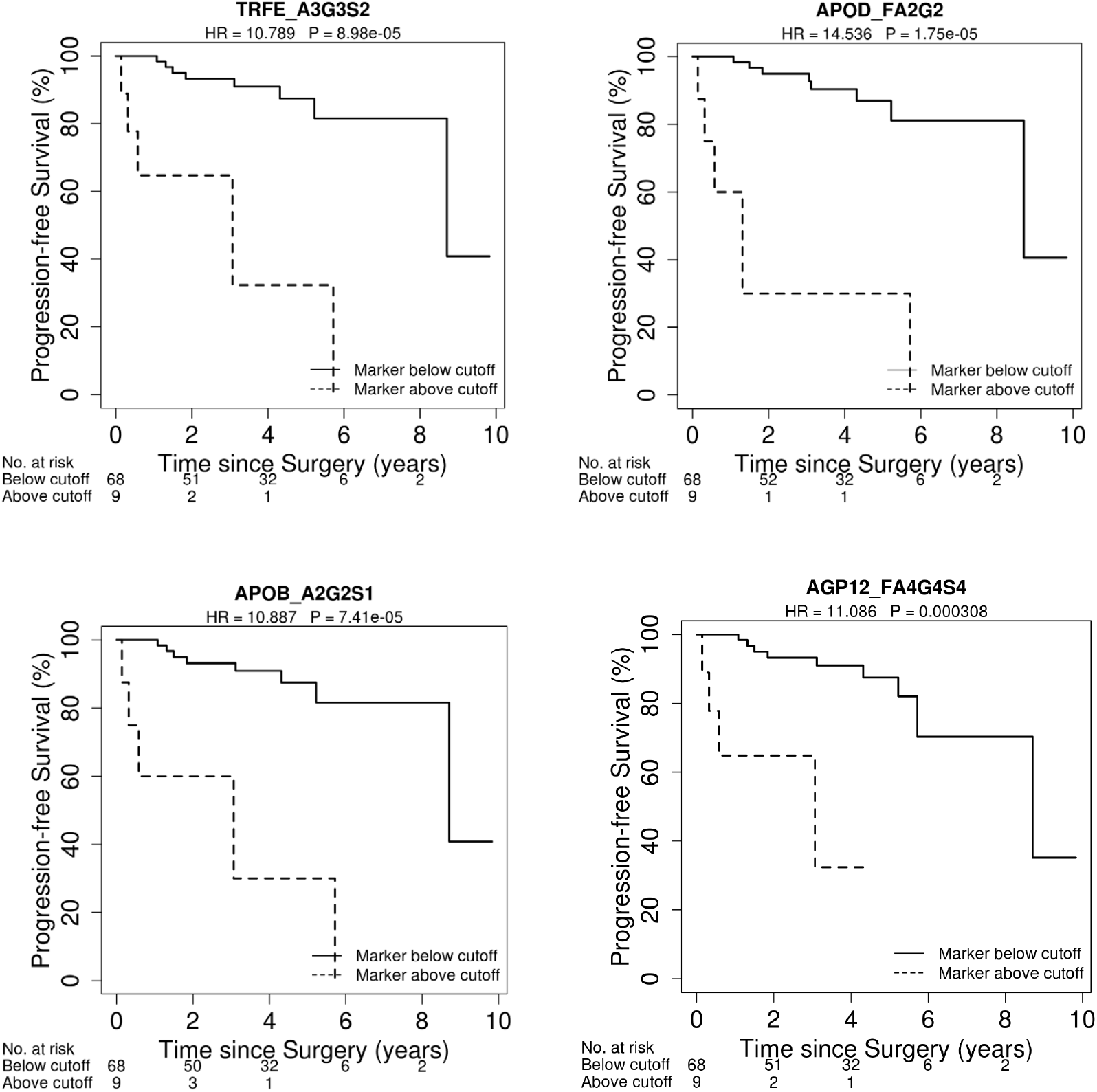
Kaplan Meier curve of glycoprotein markers with dichotomized HR > 10

Glycopeptides with continuous HR > 6 and FDR < 0.05 were utilized for multivariable model building. Three markers (prothrombin A2G2S, immunoglobulin J chain G2FS, and complement component C8A A2G2S2) were retained via backwards selection. The three markers were used to generate multivariable LOOCV prediction models to determine progression. Predicted scores were dichotomized at cutoff that maximized Harrell’s c-index and resulting Kaplan-Meier curve is plotted in Figure 3a (HR = 11.96, p = <0.001). Stratification by tumor stage demonstrates applicability in low stage (Figure 3b, HR = 14.56, p = 0.001) and high stage tumors (Figure 3c, HR =9.72, p = 0.057). One patient was not included in stratification by tumor stage due to missing stage.

**Figure 3a.**
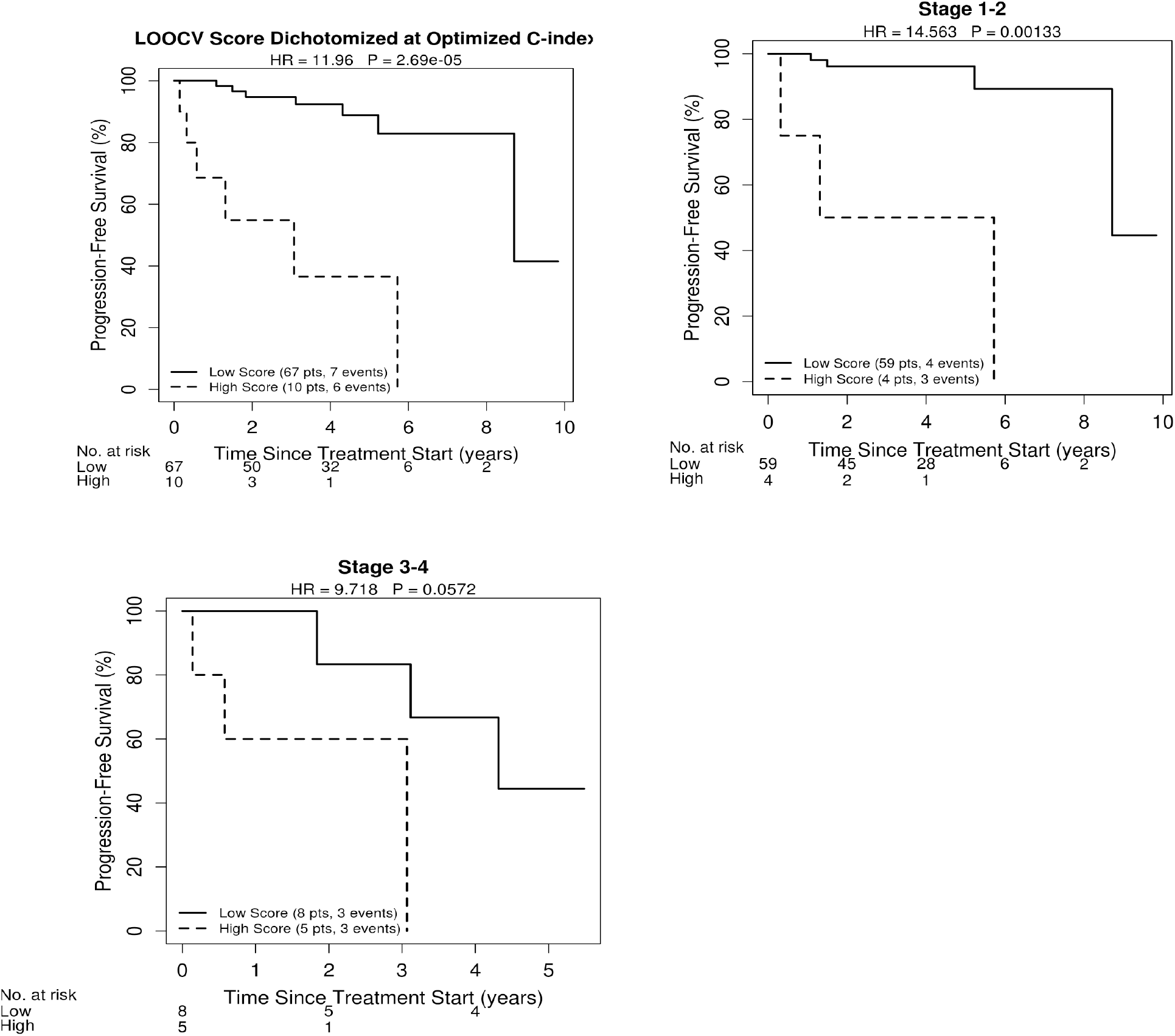
Kaplan-Meier (KM) curve of predicted survival score generated via LOOCV, utilizing prothrombin A2G2S, immunoglobulin J chain FA2G2S2, and complement component C8A A2G2S2. Figure 3b. KM curve stratified by stages 1-2. Figure 3c. KM curve stratified by stages 3-4.

To further assess biological significance of the biomarkers identified and to investigate their putative association with cancer biology, these biomarkers were subjected to pathway analysis to identify canonical pathways, protein networks, and predicted upstream regulators of relevance. The 10 statically most significantly enriched pathways with an overlapping *p*-value of < 0.01are shown in Figure 4A. Three candidate biomarkers, APOM, CLU, and F2, are associated with the clathrin-mediated endocytosis signaling pathway. APOM and CLU are involved in the liver X receptor and retinoid acid X (LXR/RXR) pathway as well. The five statically most significantly enriched pathways also include the FXR/RXR pathway, atherosclerosis signaling, and IL-12 signaling and production in macrophages. Upstream regulators analysis identified three transcription factors, HNF1A, JUNB and TAF12, at *p* values of <0.001 (Figure 4B). Among them, JunB was reported to modulate tumor invasion and angiogenesis of ccRCC through matrix metalloproteinase-2 (MMP-2), MMP-9 and chemokine (C-C motif) ligand-2 (CCL2)[11]. A GWAS study of the human N-glycome has shown that HNF1α acts as a master regulator of plasma protein fucosylation through regulating the expression of major fucosyltranferase and fucose biosynthesis genes [12]. Proteins interacting directly or indirectly with the statistically significant glycopeptides in our study are consolidated into the network. The network is overlaid with functions and presented as a graph indicating the molecular relationships between proteins (Figure 4C). The results suggest that 24 (including C8A, JCHAIN, F2 and CLU) of 31 proteins in the network are associated with cancer development and progression.

**Figure 4.**
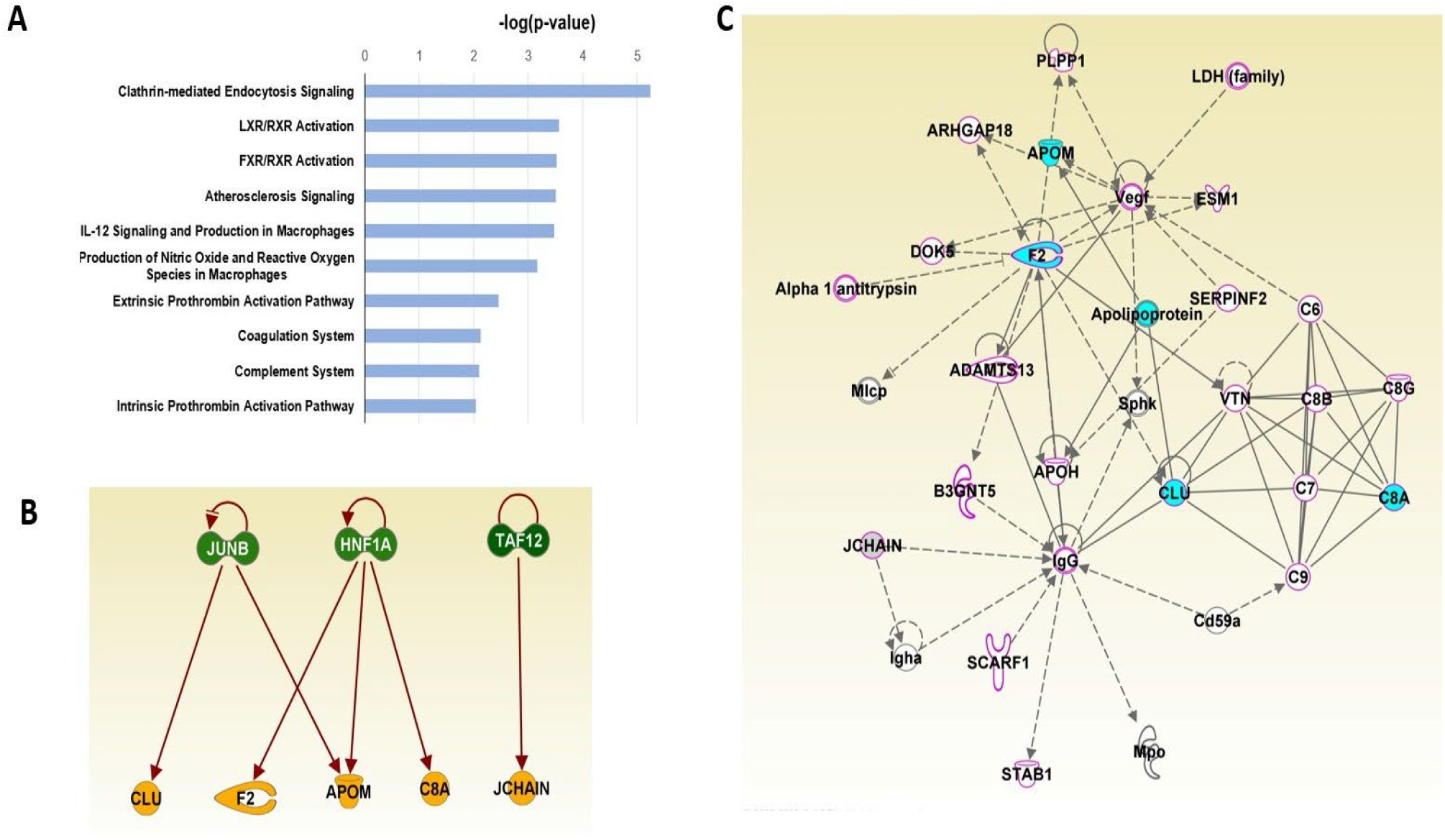
IPA analysis of candidate biomarkers. (A) Top 10 canonical pathways enriched with candidate biomarkers (*P* <0.01). (B) Top 3 upstream transcription factor regulators predicted. (C) Molecular protein network generated in IPA. The network is overlaid with disease and functions. The nodes highlighted with cyan are the candidate biomarkers. The molecules outlined with magenta are involved in cancer development based on knowledge findings. Solid lines represent direct interactions between two proteins and dotted lines show indirect interactions.

## Discussion

Protein glycosylation characteristics are affected by structural and regulatory modulations at the DNA and RNA level of enzymes, as well as by metabolic, inflammatory, and immunologic changes within the intra- and extracellular microenvironment. All of these have been proposed as factors in carcinogenesis. Changes of protein glycosylation may be envisioned as uniquely informative biomarkers as they represent the ultimate downstream product of cellular anabolism. In the case of ccRCC, downregulation of oxidative phosphorylation was recently demonstrated without observed changes at the RNA level, indicating that identifying drivers of oncogenesis must include downstream regulators, including evaluation of PTM[13].

Despite the evidence of PTM essential role in oncogenesis, biomarkers have been limited by the challenges associated with analysis. These are a complexly heterogeneous class of molecules that are difficult to characterize. Determining their specific localization within a given protein backbone represents an additional layer of challenges. Thus, the commonly used approach towards glycoproteomic characterization, namely the quantification in aggregate of all individual glycans released by enzymatic treatment of one or more proteins, ignores site-specificity aspects of glycosylation and materially abandons the most critical information about the modification [6]. Likewise, studies that interrogate protein glycosylation sites in the absence of glycan information -- due to glycan degradation-- are not capturing the depth of information present [14]. Other approaches, such as depletion of abundant proteins to improve analysis of lower abundance proteins are similarly limiting for many reasons [1]. The site-specific glycoprotein analysis developed by Li et al. allows rapid, analysis of integrated proteomic and glycan data [7], without the bias introduced by depletion or enrichment. This has dramatically improved quantification efficiency of N-glycans in serum [5] and with use of advanced software-enhanced platform has fundamentally changed the scalability.

The results of our study provide, for the first time to our knowledge, a set of biomarkers that highly correlates with prognosis and disease course of individuals diagnosed with ccRCC and treated with surgical tumor resection. We identified 36 glycosylated proteins in the serum of ccRCC patients that exhibited statistically significant expression differences in accordance with subsequent disease course. Five of these markers demonstrated hazard ratios above 3.5 (from 3.9-10.7) for PFS.

Most early studies of glycosylated protein markers have focused on early diagnosis and screening with limited evaluation of prognostication. A recent study in ovarian cancer demonstrated differences in the global glycoproteome as well as site specific differences between benign and malignant ovarian tumors[15], highlighting the importance of quantifying site-specific changes that would be otherwise missed. Likewise, site-specific glycosylation changes demonstrated in liver cirrhosis may be helpful as screening tools for hepatocellular carcinoma [16, 17].

Glycosylated protein biomarkers are still in their infancy. Renal cell carcinoma is a heterogeneous disease with multiple molecular pathways leading to differential tumorigenesis. External validation on a larger cohort is essential given the multiple subsets of proteomic alterations identified in ccRCC[13]. The major limitation of this study is the relatively large number of individuals lost to follow-up, impacting the applicability to larger cohorts. Also, the underrepresentation of more advanced-stage disease limits this subset. Another challenge is the lack of functional insights into the nature of the glycoprotein-isoform differences associated with phenotype. To our knowledge, none of the glycoproteins identified have been previously associated with ccRCC biology. It is unclear whether these glycopeptides are byproducts of the tumor released from the cell surface by matrix metalloproteases or whether they represent a more systemic inflammatory or immunologic reaction to the tumor. Clearly, additional experimentation will be needed to interrogate these questions. However robust association of glycoprotein profiles with tumor phenotype, if confirmed, can be a valuable tool for prognostication and management of patients. Thus, glycoprotein profiling may help identify patients who may benefit from closer surveillance or adjuvant therapy after surgical treatment for ccRCC. Additionally, this approach may be explored in characterizing small renal masses which currently represents another diagnostic challenge.

## Conclusions

Glycoprotein isoform differences measured in serum samples of patients with ccRCC prior to undergoing surgical treatment were found to be strongly associated with subsequent disease course and tumor recurrence. Our results warrant further studies to examine the findings more closely, and in larger cohorts. If confirmed, this may open the door for a more targeted, and potentially more effective, approach to the management of ccRCC.

## Data Availability

All data produced in the present study are available upon reasonable request to the authors

**Appendix A.**
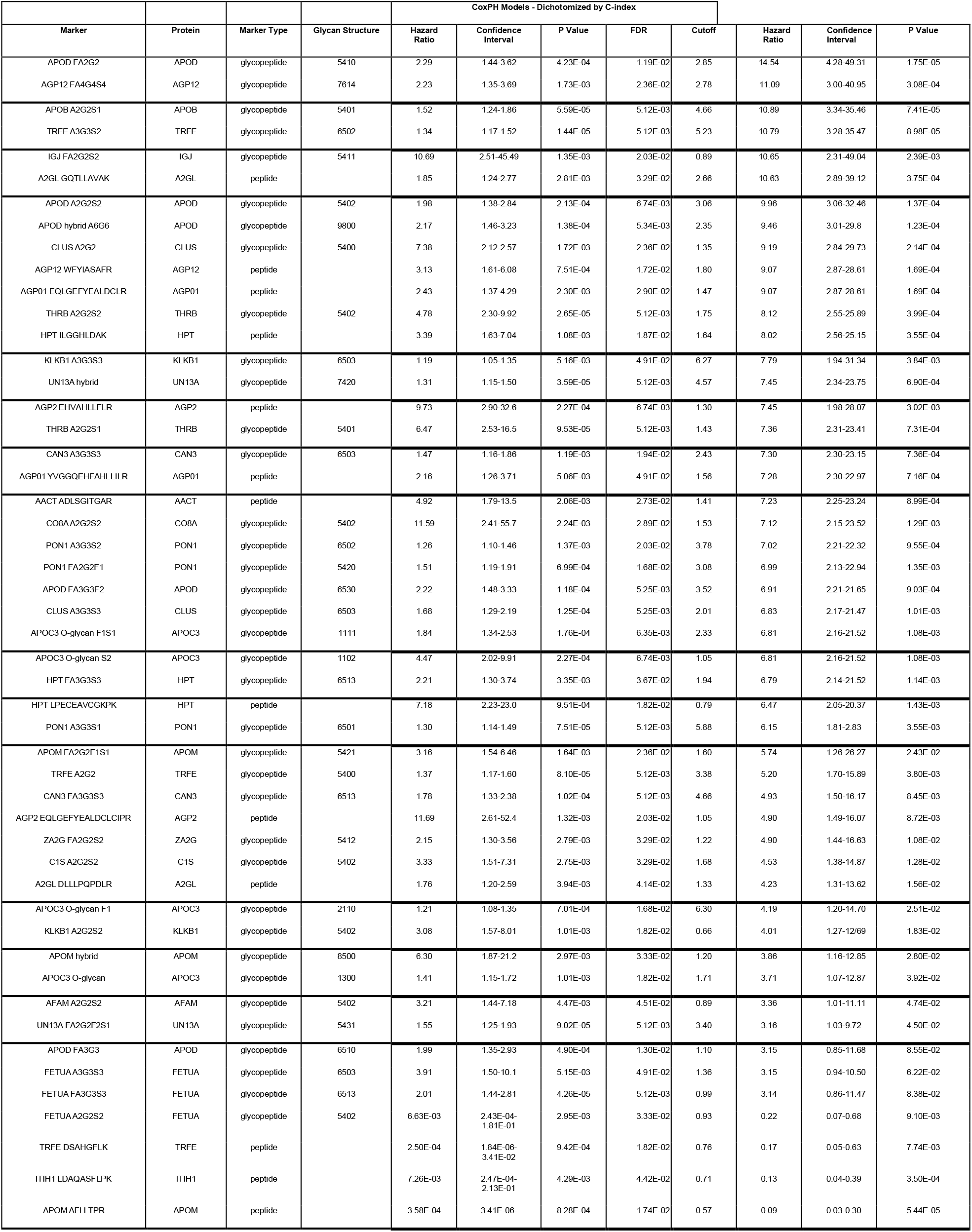
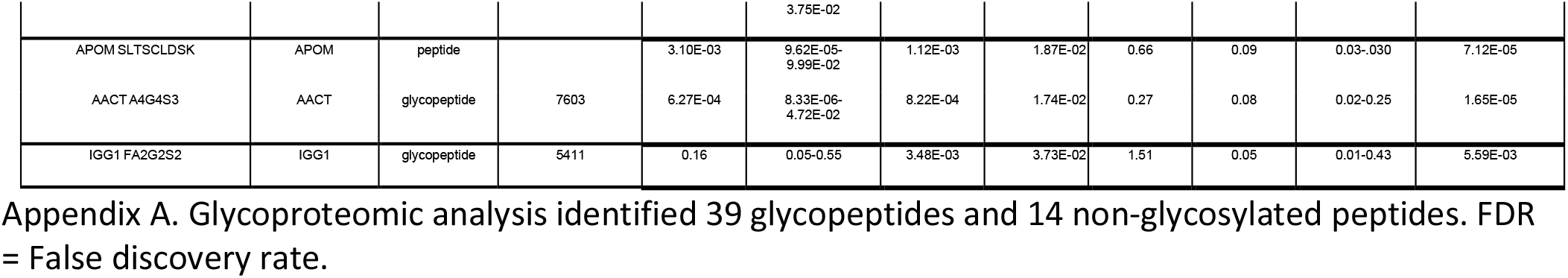
Glycoproteomic analysis identified 39 glycopeptides and 14 non-glycosylated peptides. FDR = False discovery rate.

**Appendix B.**
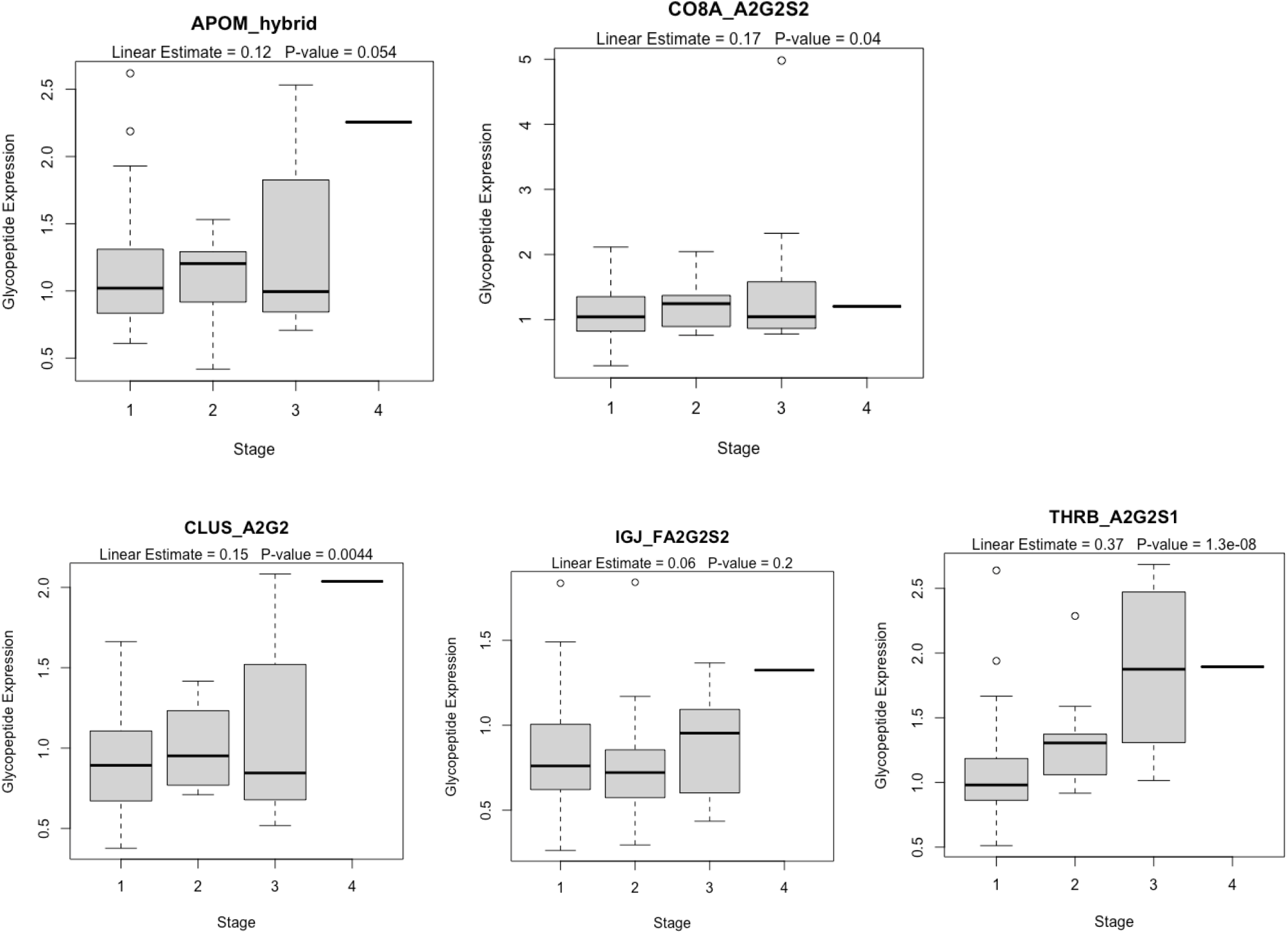
Linear Regression Analysis of Association of the five glycoprotein markers with continuous hazard ratio >6 with increasing tumor stage.

## References

1. Lane, B.R. and M.W. Kattan, Prognostic models and algorithms in renal cell carcinoma. Urol Clin North Am, 2008. 35(4): p. 613-25; vii.

2. Iafolla, M.A.J., et al., Systematic review and REMARK scoring of renal cell carcinoma prognostic circulating biomarker manuscripts. PLOS ONE, 2019. 14(10): p. e0222359.

3. Pinho, S.S. and C.A. Reis, Glycosylation in cancer: mechanisms and clinical implications. Nat Rev Cancer, 2015. 15(9): p. 540–55.

4. Drake, P.M., et al., Sweetening the Pot: Adding Glycosylation to the Biomarker Discovery Equation. Clinical Chemistry, 2010. 56(2): p. 223–236.

5. Alley, W.R., et al., N-linked Glycan Structures and Their Expressions Change in the Blood Sera of Ovarian Cancer Patients. Journal of Proteome Research, 2012. 11(4): p. 2282–2300.

6. de Leoz, M.L., et al., Glycomic approach for potential biomarkers on prostate cancer: profiling of N-linked glycans in human sera and pRNS cell lines. Dis Markers, 2008. 25(4-5): p. 243–58.

7. Li, Q., et al., Site-Specific Glycosylation Quantitation of 50 Serum Glycoproteins Enhanced by Predictive Glycopeptidomics for Improved Disease Biomarker Discovery. Anal Chem, 2019. 91(8): p. 5433–5445.

8. Jorns, J.J., et al., Kidney size and cancer-specific survival for patients undergoing nephrectomy for pT1 clear cell renal cell carcinoma. Urology, 2012. 80(1): p. 147–50.

9. Wu, Z., et al., PB-Net: Automatic peak integration by sequential deep learning for multiple reaction monitoring. J Proteomics, 2020. 223: p. 103820.

10. Benjamini, Y. and Y. Hochberg, Controlling the False Discovery Rate: A Practical and Powerful Approach to Multiple Testing. Journal of the Royal Statistical Society: Series B (Methodological), 1995. 57(1): p. 289–300.

11. Kanno, T., et al., JunB promotes cell invasion and angiogenesis in VHL-defective renal cell carcinoma. Oncogene, 2012. 31(25): p. 3098–110.

12. Lauc, G., et al., Genomics meets glycomics-the first GWAS study of human N-Glycome identifies HNF1α as a master regulator of plasma protein fucosylation. PLoS Genet, 2010. 6(12): p. e1001256.

13. Clark, D.J., et al., Integrated Proteogenomic Characterization of Clear Cell Renal Cell Carcinoma. Cell, 2019. 179(4): p. 964-983.e31.

14. Zeng, X., et al., Lung cancer serum biomarker discovery using glycoprotein capture and liquid chromatography mass spectrometry. J Proteome Res, 2010. 9(12): p. 6440–9.

15. Li, Q.K., et al., An integrated proteomic and glycoproteomic approach uncovers differences in glycosylation occupancy from benign and malignant epithelial ovarian tumors. Clinical Proteomics, 2017. 14(1).

16. Lee, H.J., et al., Abundance-ratio-based semiquantitative analysis of site-specific N-linked glycopeptides present in the plasma of hepatocellular carcinoma patients. J Proteome Res, 2014. 13(5): p. 2328–38.

17. Zhu, J., et al., Glycoproteomic markers of hepatocellular carcinoma-mass spectrometry based approaches. Mass Spectrom Rev, 2019. 38(3): p. 265–290.

